# A European clinical practice guideline on pain, agitation, delirium, and iatrogenic withdrawal syndrome management in critically ill children: A protocol

**DOI:** 10.1101/2024.10.12.24315389

**Authors:** Ibo MacDonald, Alexia Cavin-Trombert, Cécile Jaques, Angela Amigoni, Anne-Sylvie Ramelet, the European Society of Paediatric and Neontal Intensive Care (ESPNIC) PAin Delirium Iatrogenic withdrawal Sedation (PaDIS) consortium

**Affiliations:** Institute of Higher Education and Research in Healthcare, Department of Biology and Medicine, University of Lausanne, Lausanne, Switzerland; Medical Library, Lausanne University Hospital and University of Lausanne, Lausanne, Switzerland; Pediatric Intensive Care unit, Azienda Ospedale Università Padova, Padova, Italy; Institute of Higher Education and Research in Healthcare, Department of Biology and Medicine, University of Lausanne, and the Department Woman-Mother-Child, Lausanne University Hospital, Lausanne, Switzerland

**Author notes:** These authors have contributed equally to this work and share last authorship. Corresponding author (ASR). Membership of the ESPNIC PaDIS consortium is provided in the Acknowledgements.

**Keywords:** Assessment – Pediatric intensive care – Treatment – Critical care – Comfort

## Abstract

**Introduction:** In pediatric intensive care units, pain, sedation, delirium, and iatrogenic withdrawal syndrome must be managed as interrelated conditions. Existing clinical practice guidelines have some methodological limitations and are not readily transferrable to the European context without adaptation. This protocol describes the methods for developing a high-quality, and the first patient- and family-informed European guideline for managing pain, sedation, delirium and iatrogenic withdrawal syndrome.

**Methods:** The guideline will be developed using the Grading of Recommendations Assessment, Development and Evaluation (GRADE) - ADOLOPEMENT approach, engaging clinical experts and patients and families in the development process. It will consist of seven phases: 1) Set-up – establishing three groups for guideline development: i) steering committee, ii) development panel (experts and patient/family partners from across Europe), and iii) patient and family advisory panel, and scoping to determine population, conditions, purpose, and users of the guideline; 2) Preparation – voting on summary recommendations, prioritizing research questions and outcomes, and matching research questions with existing recommendations; 3) Search and retrieval of evidence – using three search approaches to develop search strategies to find evidence, conducting individualized searches for each summary recommendation and new research question, selecting evidence, study appraisal, and initial data extraction; 4) Evidence synthesis – summarizing evidence in evidence profiles and summary of evidence tables, and conducting panel surveys of current practice when evidence is absent; 5) Development – drafting recommendations, voting on and approving them, and developing accompanying materials; 6) Review – conducting internal, society-level, and external international expert reviews; and 7) Issue – publishing the guideline.

**Discussion:** This protocol ensures a transparent process follows the GRADE approach for guideline development, leading to a high-quality, trustworthy, and credible guideline for managing pain, sedation, delirium and iatrogenic withdrawal syndrome in critically ill children. Tailored to the European context for healthcare professionals.

**Registration:** Practice guideline REgistration for transPAREncy (PREPARE) registration number PREPARE-2024CN859

## Introduction

Analgesia and sedation in critically ill children remain challenging for healthcare professionals in pediatric intensive care units (PICUs), due to the heterogeneous responses of patients to the same medications and the potential for drug side effects [1]. Suboptimal sedation can cause discomfort, agitation and increased medication use [2]. While oversedation may cause complications like diaphragm dysfunction, hemodynamic instability, altered bowel function, increased morbidity risk, and prolonged PICU stay [2, 3]. Undersedation can increase anxiety, stress, and the risk of accidental medical equipment removal, including unplanned extubation [2, 4]. Prolonged use of analgesics and sedatives is associated with iatrogenic withdrawal syndrome (IWS) and delirium [5, 6], both having long-term cognitive, emotional, and social impairment in children, and causing significant distress for families [7, 8]. Therefore, achieving optimal analgosedation levels in critically ill children is fundamental for their comfort and safety with close monitoring.

Accurate assessment is a prerequisite for appropriate management. The 2016 European Society of Pediatric and Neonatal Intensive Care (ESPNIC) recommendations emphasized the importance of using validated measurement instruments to monitor pain, depth of sedation, delirium, and IWS [9]. However, recent surveys have highlighted wide variations in assessment practices and choice of drugs for managing pain, sedation, delirium, and IWS (hereafter, referred to as the four conditions) [10–12], Additionally, compliance with these recommendations has been inconsistent, with up to 69% of clinicians not selecting the correct measurement instruments, 30% misapplying them, and 3-58% not using them at all [13], leading to potential under- or over-use of medications and adverse events.

To improve assessment and management practices, clinical practice guidelines (CPGs) have been developed to synthesize evidence into evidence-based recommendations [14]. Although two CPGs for managing pain, sedation, delirium, and IWS were published in 2022, they do not incorporate newer pharmacological approaches and have outdated or flawed literature searches [15–17]. Our systematic review found variability in the quality and evidence supporting recommendations in the included CPGs. While some recommendations were supported by strong evidence, others, especially those related to delirium, lacked support [17]. Furthermore, only 22% of the CPGs (n = 4) used the recommended Grading of Recommendations Assessment, Development and Evaluation (GRADE) approach, which is important for quality CPGs [18]. Among the shortcomings in the use of GRADE in these CPGs were, limited use of PICO questions (50%), absence of prioritized outcomes (100%), and the absence of available evidence to decision (EtD) tables (100%) [17, 19]. A recent narrative review highlighted 15 gaps in current CPG recommendations that need addressing in future CPGs [20]. These limitations and emerging new evidence, underscore the need for updating and contextualizing existing evidence into a new CPG. Therefore, we aim, with the endorsement of ESPNIC, to develop a comprehensive and trustworthy CPG for managing pain, sedation, delirium, and IWS in Europe.

## Methods

The protocol for developing our CPG was registered in the Practice guideline REgistration for transPAREncy (PREPARE) under registration number PREPARE-2024CN859. The GRADE-ADOLOPMENT approach will be used for adopting, adapting, and developing de novo recommendations based on prioritized research questions and outcomes, and matching recommendations from existing CPGs [21]. This approach involves seven phases, ten steps, and 22 actions, summarised in Fig 1 and described below. As no reporting checklist exists for guidelines, one was developed using Xun et al. [22] (S1 File).

**Fig 1:**
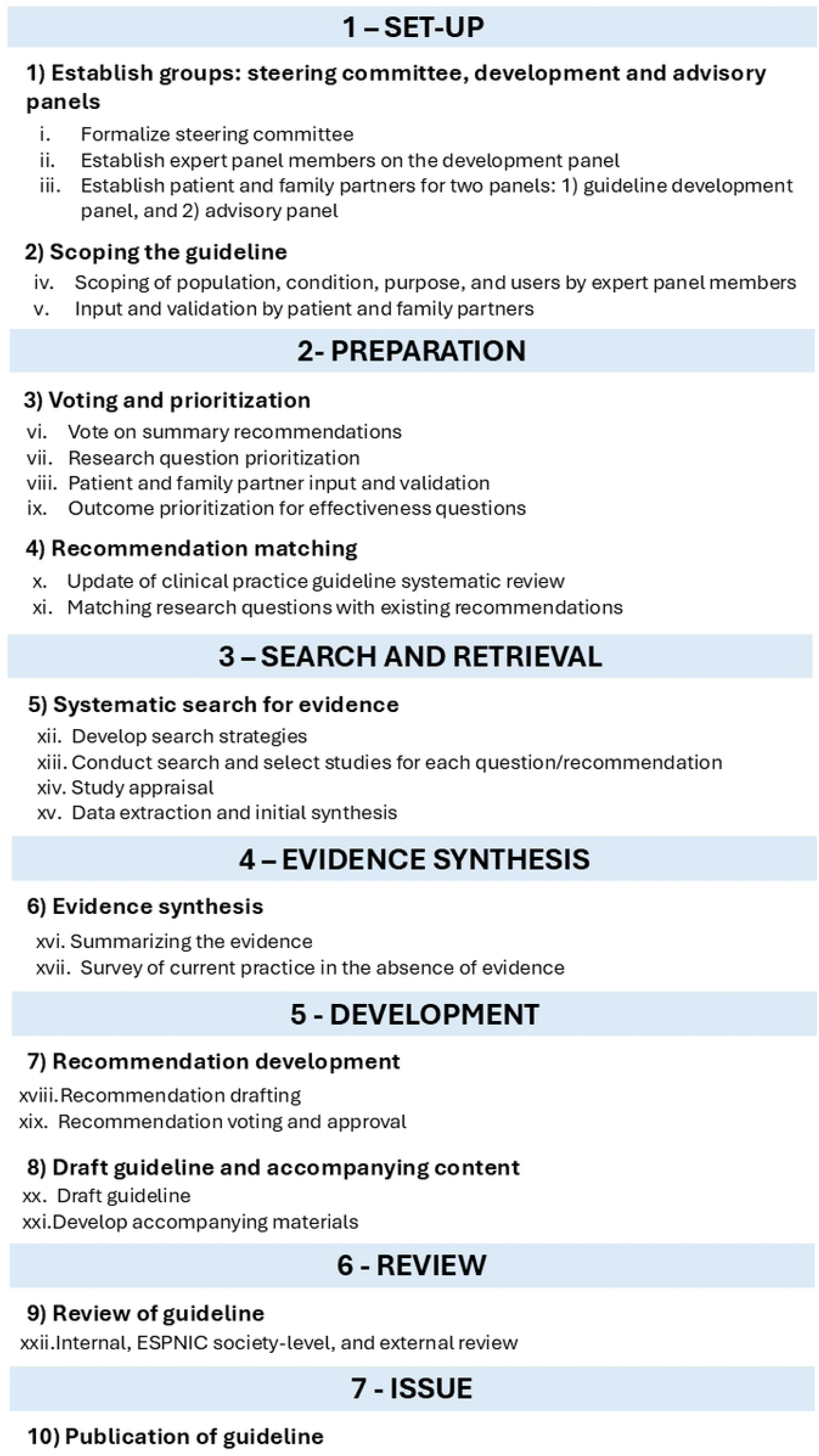
Phases, steps, and actions of the guideline development process.

## Phase 1: Set-up

The set-up phase comprises two steps and five actions.

### Step 1: Establish groups: steering committee, development and advisory panels

Three groups will be formed: the steering committee, the development panel, and the patient and family partner (PFP) advisory panel.

#### Action 1 – Formalize the steering committee

The steering committee includes a pediatric intensivist (AA), a PICU nurse and researcher (ASR), and a nurse methodologist (IMD). The steering committee will oversee, organize, and coordinate the development process. Committee members (ASR, AA) are institution-supported and receive no funding for their work, while the chair (IMD) is supported by a postdoctoral fellowship for her role and will also take on all administrative support tasks. An invited guest to the steering committee will be the lead health sciences information specialist (ACT), who will assist with planning and conducting the literature reviews.

#### Action 2 – Establish development panel

The development panel will include clinical experts and Patient and Families Partners.

*<u>A. Clinical expert members.</u>* In April 2023, a call for expressions of interest to join the development panel was sent to the Analgosedation Consortium from the Pharmacology and the Nursing Science Sections of ESPNIC. Interested members completed a declaration of interest and conflict of interest (COI) form (S2 File). However, recruitment is ongoing. The expert panel will include nurses, intensivists, and pharmacists from as many European countries as possible, all of whom will volunteer their time.
*<u>B. PFP members</u>.* PFPs who have specific experience with one or more aspects of the CPG will be involved and engaged in the different steps of the development process as development panel members. Recruitment of all PFPs is explained in Action 3.

#### Action 3 – Patient and Family involvement and engagement

Expert panel members will identify families and/or patients from their existing networks and provide them with an information sheet (File S3) and an expression of interest form that includes COI (File S4) [23–27]. If translation of these forms is required, an established translation method will be used by the bilingual native-speaking expert panel member [28].

Interested patients and family members will be able to choose to join either the development panel or the PFP advisory panel, depending on their level of comfort with English and willingness to engage. They will indicate their group preference on the expression of interest form.

PFPs on the development panel will be compensated at an hourly rate for their participation in panel meetings. For tasks outside of meetings, such as voting and reviewing, all PFPs will receive compensation on a flat rate basis. The compensation was determined by averaging the compensation rates used by represented countries and by referring to recommendations from the National Institute for Health and Care Research [29]. PFP compensation is provided by ESPNIC.

The roles, responsibilities, and level of engagement and involvement across the three groups are outlined in Table 1

**Table 1:**
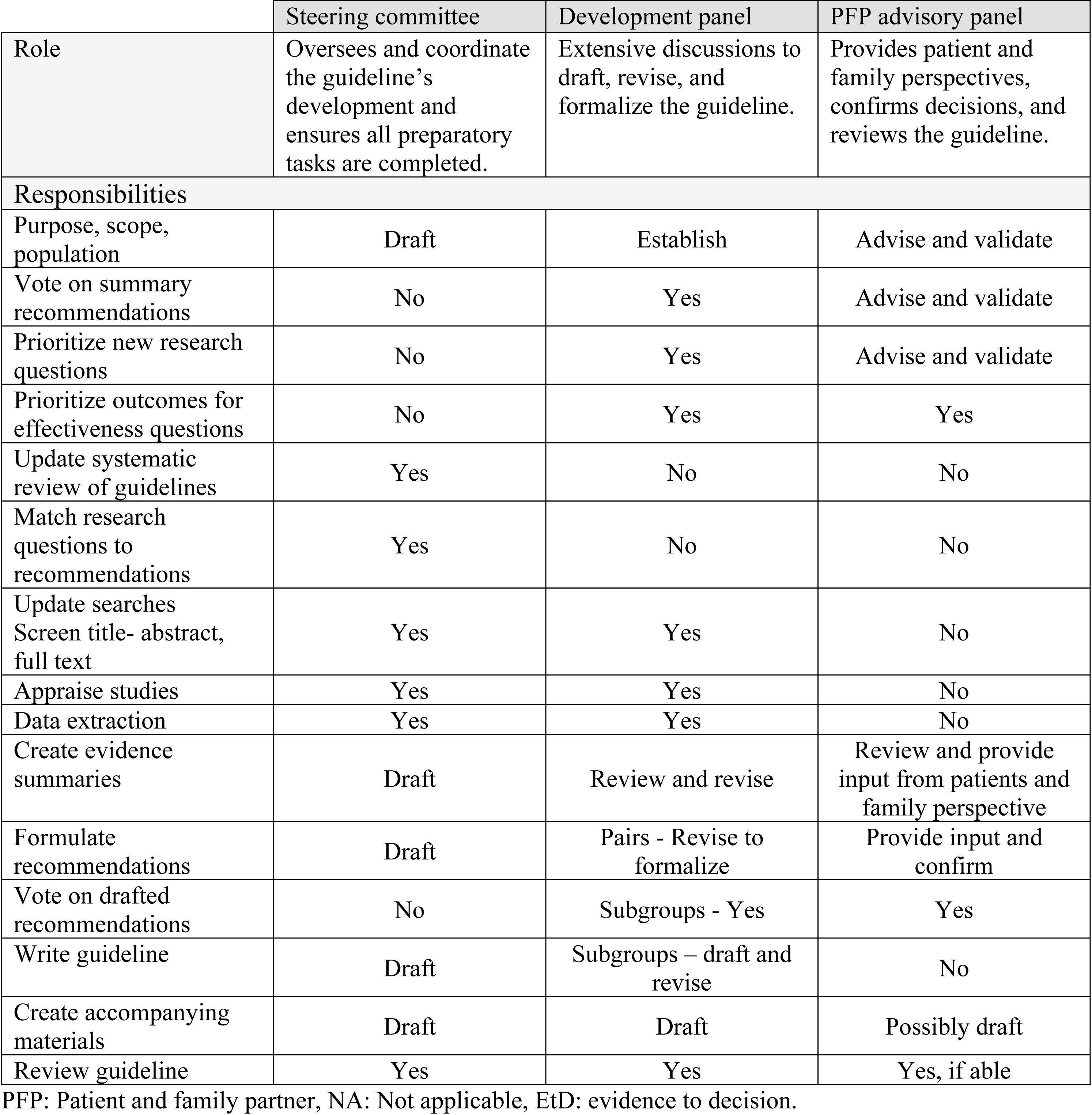
Roles and responsibilities of the three groups.

|  | Steering committee | Development panel | PFP advisory panel |
| --- | --- | --- | --- |
| Role | Oversees and coordinate the guideline’s development and ensures all preparatory tasks are completed. | Extensive discussions to draft, revise, and formalize the guideline. | Provides patient and family perspectives, confirms decisions, and reviews the guideline. |
| Responsibilities |  |  |  |
| Purpose, scope, population | Draft | Establish | Advise and validate |
| Vote on summary recommendations | No | Yes | Advise and validate |
| Prioritize new research questions | No | Yes | Advise and validate |
| Prioritize outcomes for effectiveness questions | No | Yes | Yes |
| Update systematic review of guidelines | Yes | No | No |
| Match research questions to recommendations | Yes | No | No |
| Update searches<br>Screen title- abstract, full text | Yes | Yes | No |
| Appraise studies | Yes | Yes | No |
| Data extraction | Yes | Yes | No |
| Create evidence summaries | Draft | Review and revise | Review and provide input from patients and family perspective |
| Formulate recommendations | Draft | Pairs - Revise to formalize | Provide input and confirm |
| Vote on drafted recommendations | No | Subgroups - Yes | Yes |
| Write guideline | Draft | Subgroups – draft and revise | No |
| Create accompanying materials | Draft | Draft | Possibly draft |
| Review guideline | Yes | Yes | Yes, if able |

PFP: Patient and family partner, NA: Not applicable, EtD: evidence to decision.

All members of the development panel will complete a COI form covering financial, academic, clinical, and community aspects [30]. At the beginning of each development panel meeting, the steering committee chair will ask if any member have changes to their COI to declare. If yes, a new COI form will be completed. The steering committee will monitor COIs, members with a declared COI will not participate in the development or voting on recommendations related their COI.

Ethical approval will not be sought as no direct data from patients or clinicians will be used; instead, their experiences will be used to inform the guideline. Completion of the expression of interest form will signify consent to participate in the development of the guideline. If children are interested in participating in either panel, their parents will also complete an expression of interest form and will participate alongside them.

Training will be provided either during panel meetings or through available online resources [31–33].

### Step 2: Scoping the guideline

#### Action 4 – Scoping

The population, condition, purpose, and users of the CPG will be determined during the first online meeting with the expert panel members of the development panel.

#### Action 5 – Input and validation

Results of Action 4 will be sent to all PFPs for review, input, and validation via SurveyMonkey [34]. PFPs will indicate whether they “reject”, “accept”, or “accept with modifications” each element, providing rationale or indicating desired changes. A percentage for acceptance or rejection will be calculated for each element, with a consensus of 80% or more required for confirmation. The steering committee will review and revise as needed and will present these at the next development panel meeting.

## Phase 2: Preparation

The preparation phase includes two steps and six actions.

### Step 3: Voting and prioritization

#### Action 6 – Voting on summary recommendations

The 30 summary recommendations synthesized in our systematic review of CPGs [17] were chosen for voting because they were featured across multiple CPGs. Additionally, their supporting evidence was reviewed, along with harmonizing their strength of recommendations and certainty of evidence [17]. These summary recommendations will be added to SurveyMonkey [34], with an open-ended question asking about clinical areas where recommendations are needed or lacking (see Action 7 below). Each member will select to either “accept (adopt)”, “reject”, or “accept with modifications (adapt)” each recommendation. If modifications are needed, members will be asked to specify them. Responses will be tallied, with “accept” and “accept with modifications” being grouped together. A consensus of 80% or more will be needed for inclusion. Recommendations that fail to meet this threshold will be discussed during a consensus meeting with the development panel, where a decision will be made.

#### Action 7 – Research question prioritization

A list of research questions will be created based on the gaps identified by Mondardini and colleagues [20], and areas without recommendations from the earlier survey (Action 6). These questions will be entered into GRADEpro GDT (Guideline Development Tool) [35], an online tool that assists development panels in brainstorming, prioritizing questions and outcomes, and creating evidence to decision (EtD) tables. These EtD tables summarize evidence and present benefits, risks, and other factors to support development panels in forming recommendations [21]. The prioritization process, in GRADEpro GDT, involves the following steps:

1. Consolidation and reformulation: The steering committee will consolidate the questions by removing duplicates, and where possible, reformatting them into PICO (patient, intervention, comparator, outcome) effectiveness questions [36]. Nonactionable or descriptive questions will also be included.
2. Brainstorming: Development panel members will independently review and comment on the questions and suggest new questions if needed.
3. Second consolidation: The steering committee will refine and reformat questions from the brainstorming round to prepare for voting.
4. Voting: Development panel members will independently rate the importance of each question on a 9-point Likert-scale (1-3: non-important, 4-6: important, 7-9: critically important). GRADEpro GDT will calculate mean scores and rank the questions from highest to lowest [35].
5. Decision-making: The steering committee will review the voting results and make decisions based on the mean score for each question: reject (mean 1-3), answer (mean 7-9), or list for potential inclusion in a future CPG (mean 4-6).
6. Approval: Development panel members will review and approve the decisions made in step 5.
7. Finalize: GRADEpro GDT will generate a matrix showing the percentage approval for each question [35]. Questions with > 80% agreement will be retained.

#### Action 8 – PFP input and validation

The final list of retained summary recommendations (Action 6) and research questions (Action 7) will be sent to PFPs using SurveyMonkey [34]. They will confirm their agreement by selecting: “Yes”, “No”, or “Yes with modification”. PFPs can also propose new questions if they believe any are missing. A consensus of 80% or more is needed to retain questions and recommendations. The steering committee will review and revise based on suggested modifications, and results will be discussed during the next development panel meeting.

#### Action 9 – Outcome prioritization

This step focuses on prioritizing outcomes for effectiveness questions, excluding nonactionable/descriptive questions. This process mirrors Action 7, using GRADEpro GDT:

1. Brainstorming: The steering committee will generate a list of important outcomes for the PICU from the literature [17, 37–39]. Outcomes will be assigned to each question, and the development panel will independently review these and can suggest others.
2. Condense: The steering committee will review and consolidate outcomes as needed.
3. Voting: The development panel members will independently rate the importance of each outcome to each question using the 9-point Likert scale in GRADEpro GDT. Simultaneously, PFPs will rate the same questions and outcomes via a survey in SurveyMonkey [34] to ensure patient and family perspectives.
4. Decision-making: The steering committee will review the voting results and make decisions based on the mean score for each outcomes to: reject (mean 1-3), include in EtD table (mean 7-9), or to list the outcome in the body/text for that research question (mean 4-6). A maximum of seven outcomes can be included in EtD tables per question [40, 41]; in the event of more than seven being identified as critically important (mean 7-9), responses between experts and PFP will be compared and PFP responses will be prioritized when there are significant differences.
5. Approval: Development panel members will review and approve the decisions made in step 4.
6. Finalize: The approval matrix will be used to determine outcome inclusion, with at least 80% agreement needed to be retained.

### Step 4: Recommendation Matching

The cornerstone of the GRADE-ADOLOPMENT approach is adopting and adapting recommendations from existing CPGs by matching them to questions [21]. This process involves evaluating the relevance, applicability, and strength of the original recommendations [21].

#### Action 10 – Update of CPG systematic review

To ensure a current list of recommendations for matching with questions (Action 9), we will update our systematic review of CPGs for managing the four conditions following the same methods used previously [17].

#### Action 11 – Matching research questions with existing recommendations

The steering committee members will assess whether each prioritized question matches any existing recommendations from the six selected CPGs [9, 15, 16, 42–44] from our systematic review [21]. A match is required with the population, intervention and comparator, and at least one shared outcome. This step will help establish the searches in phase 3.

## Phase 3: Search and retrieval

### Step 5: Systematic search for evidence

This phase involves systematic searching for evidence using three distinct search approaches, based on the strength of recommendations from the summary recommendations and matched recommendations (Search approaches 1 and 2), and for unmatched or new questions (Search approach 3), as illustrated in Fig 2.

**Fig 2:**
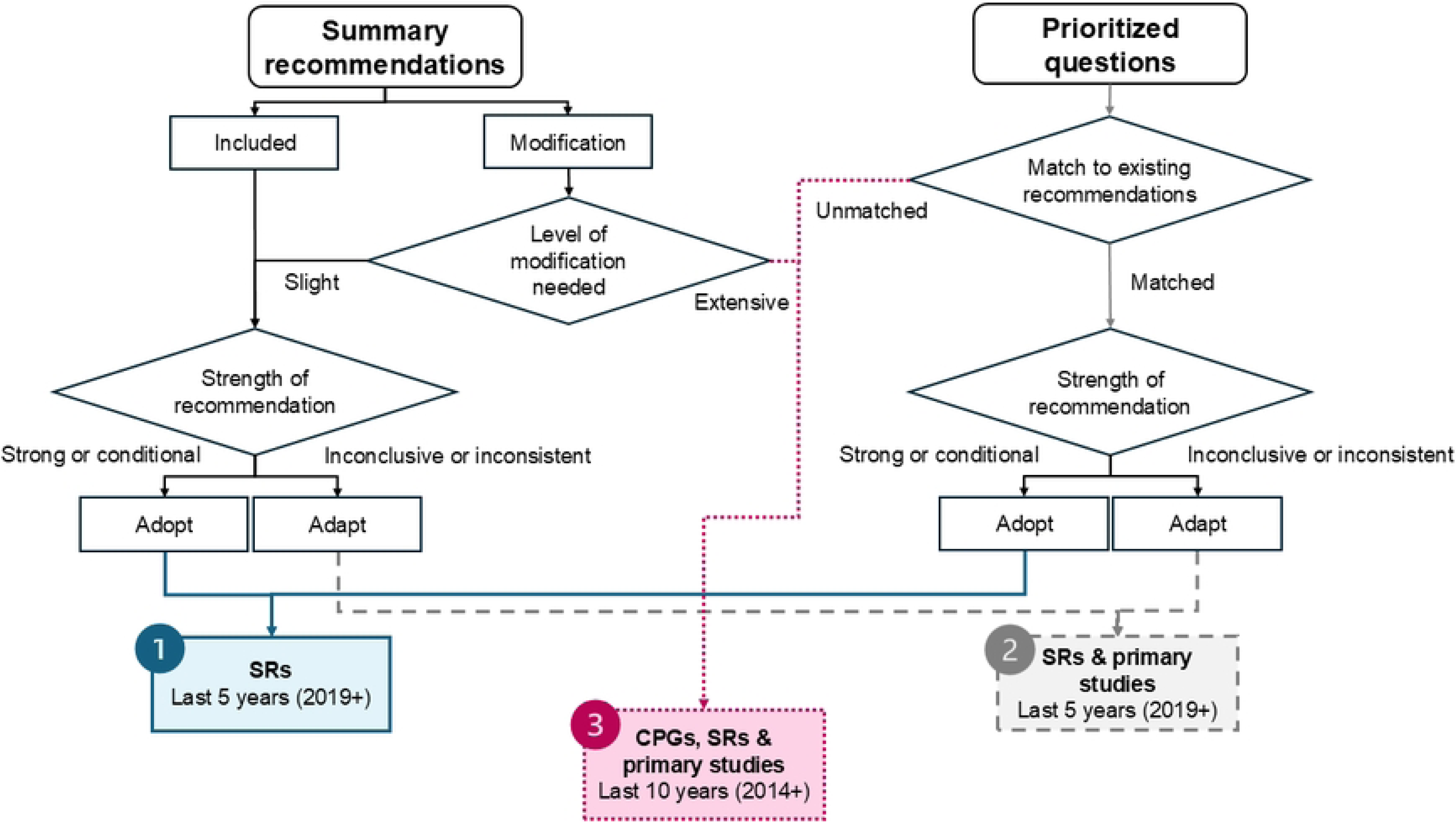
Three search approaches.

#### Action 12 – Develop search strategies

*Search approach 1:* For recommendations with a strong or conditional strength, a search for systematic reviews will be conducted.

*Search approach 2*: For recommendations with inconclusive and inconsistent strength, a search for systematic reviews and primary studies will be completed.

Both search approaches (1 and 2) will limit results to the last 5 years to ensure up-to-date evidence.

*Search approach 3:* For new and unmatched questions, or summary recommendations needing extensive modifications, a combined search for CPGs, systematic reviews, and primary studies will be conducted, with a limit of the last 10 years.

The three different search approaches are necessary to ensure that the most up-to-date evidence is incorporated into existing recommendations based on their current strength, while ensuring that all relevant evidence is retrieved for new or unmatched questions. These approaches will provide the foundation for developing comprehensive, tailored search strategies relative to study designs that help to either strengthen recommendations (Search approaches 1 and 2) or for determining the level of evidence for new questions (Search approach 3).

The search strategies for each summary recommendation and research question, developed based on one of the three search approaches, will be conducted in four electronic databases: Medline ALL (Ovid), Embase.com, CINAHL with Full Text (EBSCO), and Epistemonikos.

To update the evidence for each summary recommendation, the summary recommendation will be transformed into a research question. These questions and new questions will all have customized search strategies developed by a steering committee member (IMD) in consultation with a health sciences librarian (ACT or CJ) using index and free-terms describing the condition of interest, the population of interest (children aged 0 to 18 years), the comparison, and outcome. Another health services librarian will peer review each search strategy per question using the PRESS checklist [45].

No language restrictions will be applied, non-English studies will be translated using a standardized method [28], Deepl [46] to translate and than verification by a native speaker on the development panel or finding one via panel members’ networks. If one cannot be found, the study will not be included.

#### Action 13 – Execute search and selection

For each search, the selection process will involve the retrieved records being imported into Endnote 20 (Clarivate Analytics, USA) and duplication by using Deduklick’s automated algorithm [47]. Screening and full-text review will be independently conducted by two reviewers from either the steering committee or the development panel using Rayyan QCRI (Qatar Computing Research Institute, Doha, Qatar) [48].

Disagreements will be resolved through consensus or by a third reviewer.

#### Action 14 – Study appraisal

Appraisal will be based on study type: systematic reviews, primary studies, or CPGs, and will be conducted by two independent appraisers.

*Systematic reviews.* Quality will be assessed using A MeaSurement Tool to Assess systematic Reviews 2 (AMSTAR 2), which includes 16 items to determine overall quality ranging from critically low to high based on the presence of critical flaws [49]. Moderate and above studies will be retained.

*Primary studies.* The JBI checklists will be used to assess the quality of primary studies: Randomized control trials [50], quasi-experimental studies [51], qualitative research [52], prevalence studies [53], cohort studies [54], economic evaluations [55], diagnostic test accuracy studies [56], case reports [54], case control studies [54], and analytical cross sectional studies [54]. Each appraiser determines the level of bias for each item and an overall appraisal to include, exclude or needing more information.

*CPGs.* The AGREE II, a 23-item validated appraisal instrument will be used to evaluate the quality of CPGs 7-point Likert scale across six domains [57], scores are summed across appraisers, and if domain 3 (rigor of development) > 60% the CPG will be retained.

The literature will continue to be monitored by monthly saved searches and added, up until the development panel completes its final voting on recommendations (Action 19).

#### Action 15 – Data extraction and initial synthesis

Two members will independently extract data from each study and verify the others, using a predefined, pilot-tested Excel sheet. For effectiveness questions, outcomes will be extracted as means for potential meta-analysis if at least three studies exist with outcomes data. If median and interquartile range (IQR) are reported, Wan’s method and Excel tool will be used to transform these into means [58].

Meta-analyses will be conducted using STATA version 17 software [59]. Random-effects models applying the Sidik-Jonkman method, will be used with standard mean difference for continuous or odds ratios for dichotomous variables. The *I*^2^ test will be used to assess statistical heterogeneity, considered low if < 40%, moderate at 30% - 60%, substantial at 50% - 90%, and considerable at 75% - 100% [18]. If heterogeneity is ≥ 40%, a sensitivity analysis will be performed by removing one study at a time to assess its influencer on the overall effect size [18]. Additionally, sensitivity analysis will be performed on PICO elements, study design, or risk of bias if sufficient studies (>2) are available to explore these other sources of heterogeneity. Results will be presented as forest plots. To explore publication bias funnel plots will be used, with Egger’s test for type I error [60]. If meta-analysis is not possible, results will be described narratively.

## Phase 4: Evidence synthesis

### Step 6: Evidence synthesis

#### Action 16 – Summarizing the evidence

The GRADE approach focuses on using the best available high quality evidence, typically from systematic reviews [18]. This involves developing two types of GRADE evidence tables [19]:

1. EtD profiles: Provide information about the body of evidence and the judgements made about the quality of evidence, included as statistical results for each outcome, per question [19]. These are completed at the end of Phase 3 by the two reviewers using GRADEpro GDT. The categories addressed by these tables include: outcomes, number of studies, study design, evidence quality factors, risk, effect, and overall quality of evidence (high, moderate, low, or very low). Evidence quality is determined by reviewing five factors that can lead to downgrading (risk of bias [61], inconsistency [62], indirectness [63], imprecision [64], and publication bias [65]), and three factors that can lead to upgrading (large magnitude of effect, dose–response gradient, and when all plausible confounders increase the confidence in the effect) [18, 66, 67]. These are used to create the summary of findings (SoF) tables.
2. SoF tables: These simplified tables summarize the EtD tables without judgment details and will include information on other factors like health benefits and harms, equity, feasibility, acceptability, and cost [19, 21]. The steering committee will assign two development panel members to each question and summary recommendation based on domain of expertise to complete the SoF tables.

For nonactionable/descriptive questions, narrative evidence profiles will be created. For summary recommendations, evidence profiles will be created using both the originally cited literature and the studies identified in the updated searches.

#### Action 17 – Survey of current practice in absence of evidence

If search results for a specific question reveal little to no supporting evidence, the steering committee will develop a survey with open-ended questions to gather information on current practices of development panel members, emphasizing cases and outcomes [68, 69]. The results will be included in an EtD table and will be presented and discussed by the development panel.

## Phase 5: Development

The development phase includes two steps and four actions.

### Step 7: Recommendation development

The recommendation development process will include several steps:

#### Action 18 – recommendation drafting

1. Draft recommendations: The steering committee will draft recommendations based on the evidence profile, and will reword summary recommendations, incorporating modifications suggested during Action 6. Recommendations will be categorized as: **Strong:** Desirable effects outweigh undesirable effects, applicable to almost all patients, **Conditional**: Desirable effects probably outweigh undesirable effects, applicable to most patients but sensitive to preferences, or **Good Practice statements**: Highly relevant with benefits far outweighing harms, even with indirect supporting evidence [66]. Drafts will be sent to the pairs of panel members responsible for each specific question or recommendation (see below point 3).
2. Recommendation table creation: Each development panel pair will complete the recommendation table (File S5) for their assigned question or recommendation. The table includes the recommendation, its direction (for or against), its strength (recommend or suggest), balance of consequences, justification, considerations for subgroups (different populations or conditions), implementation considerations, monitoring and evaluation criteria, and identified research gaps. Recommendations for any accompanying documents or materials will be sent to the steering committee (i.e., resources for implementation, patient information sheet, monitoring criteria).
3. Subgroup review: The steering committee will form subgroups focused on pain, sedation, delirium, and IWS. Each subgroup includes pairs assigned to included questions and recommendations. Subgroups will meet to review, revise the evidence tables, and update the recommendation table for those that were assigned. A steering committee member will be present to address methodology questions. The goal is to reach consensus of at least 80% on the wording, direction, and strength of each recommendation. If consensus is not reached, this recommendation will be discussed at the next development panel meeting.

#### Action 19 – recommendations voting and approval

1. Recommendation approval voting #1: The recommendations and related tables (EtD, SoF, and recommendation) will be sent to all development panel members for review at least two weeks before the next panel meeting. During the meeting, members will vote to “accept”, “reject”, or “accept with modifications”. At least 80% of panel members must be present to commence voting, with an 80% consensus required to approve a recommendation.

Recommendations not reaching 80% approval will require further subgroup revision and voting until 80% approval is achieved.

1. Recommendation review by PFP advisory panel: The final list of approved recommendations will be sent to the PFP advisory panel using SurveyMonkey for review and comment. They will be asked to “accept” or “suggest modifications” for each recommendation.
2. Recommendation finalization: Any suggested modifications will be discussed at the next scheduled development panel meeting, followed by a final round of voting to approve and finalize recommendations.

### Step 8: Drafting CPG and accompanying content

#### Action 20 – Draft guideline

Once recommendations are finalized, each recommendation pair will draft their section for the CPG. This will then be reviewed by the entire subcommittee before sending to the steering committee. The steering committee will compile, standardize, and expand (if needed) each recommendation.

#### Action 21 – Develop accompanying materials

The steering committee will convene to review all suggestions for accompanying materials from panel pairs (Action 17, step 2) and make decisions on what is needed and feasible. The committee will conduct an initial search for existing materials. Those materials that are deemed necessary will be brought to a development panel meeting for discussion and consensus, and distributed to panel members for development, adaptations, or for seeking permission to use existing materials.

## Phase 6: Review of CPG

### Step 9: Review of CPG

#### Action 22 – Review of CPG

The draft CPG will undergo three separate and sequential reviews: 1) internal review by the development panel and the PFP advisory panel, 2) consultation open to all ESPNIC members, and 3) external review by invited international experts, identified by the steering committee and development panel.

The reviews will be conducted through an anonymous online survey using SurveyMonkey [34], with a link sent by email from the steering committee chair using the ESPNIC’s internal distribution list and personal contacts. Reviewers will evaluate the guideline for overall agreement and clarity, by section, using a 9-point Likert scale. Each section will have a comments area for missing information, suggestions for modifications, setting-specific issues, and implementation implications.

The steering committee members will compile feedback from each review stage and revise the draft accordingly. The changes by review will be documented and appended to the final CPG. These will be discussed at a development panel meeting, with changes requiring 80% approval.

## Phase 7: Issue

### Step 10: Publication of guideline

The CPG development process will culminate in its publication in a peer-reviewed journal. The outcomes from phases 1 - 4 will be published in a separate publication. The Right-Ad@pt checklist [70] will be used to ensure the reporting of the adoption process. The target date for completing this CPG is June 2025. This CPG will be updated in five years or sooner if new evidence significantly impacts a recommendation as monitored by the chair (IMD) using the search alerts.

## Discussion

This protocol outlines a comprehensive methodological approach, using GRADE, to develop, adapt, and contextualize a CPG for managing pain, sedation, delirium and IWS in critically ill children. By addressing existing gaps in clinical knowledge and practice, the final CPG will provide updated, evidence-based recommendations that incorporate emerging research and clinical advancements. The rigorous methods include question and outcome prioritization, the use of the GRADE framework for evaluating evidence quality and the strength of recommendations, and the creation of EtD tables. These EtD tables will be made readily available to assist those in the interpretation of the CPG, support future CPG developers or adaptors, and aid those responsible for updating this CPG. The methods used will ensure the creation of a high-quality, transparent, and accessible CPG that will promote consistent, standardized care and improve outcomes in PICUs.

This protocol describes the approach to GRADE-ADOLOPMENT when EtD tables are unavailable, offering a step-by-step process for those adopting this method for other CPG development and/or adaptation. A key strength of this CPG is the inclusion of PFPs early and throughout all phases of the CPG development process, addressing a notable limitation in previous CPGs [17]. This patient- and family-centered approach considers a broad range of PFP experiences, enhancing the relevance and applicability of the CPG. Additionally, the involvement of a wide range of experts and PFPs from across Europe ensures representativeness and contextual sensitivity. The collaborative approach with continuous consensus meetings aims to promote alignment and buy-in from all development panel members. The methodology includes the addition of voting on summary recommendation, a novel approach to synthesizing recommendations, as well as voting on research question and prioritizing outcomes, which addresses methods previously missed in existing CPGs according to the GRADE approach. By including research question voting based on clinical gaps identified both the literature and by the experts, this process helps to identify key research areas for future investigation. A major strength is the comprehensive search approaches and tailored search strategies to each recommendation and question. While other CPGs relied on a single strategy to locate relevant literature and then distribute amongst questions [15–17], this approach employs individual strategies to ensure thoroughness and transparency, even if it means encountering some redundancy in search results.

There are several limitations, firstly, achieving consensus may be challenging due to diverse perspectives across many stakeholders and across different languages. To facilitate consensus, surveys and meetings will be used to gather input. Language barriers will be addressed by offering translated materials, providing subtitles during Zoom meetings, and ensuring a bilingual speaker is available for direct translation when necessary, in meetings. Another limitation inherent to the current state of knowledge is the availability and quality of evidence for certain conditions or interventions, which may be limited. However, consolidating this information in a transparent and accessible manner, combined with surveys from a large development panel, will assist the formulation of credible recommendations. Additionally, using standardized translation method for published studies in other languages may introduce new evidence previously overlooked.

## Conclusion

This protocol outlines a comprehensive approach to developing a CPG for managing pain, sedation, delirium and IWS in pediatric critically ill patients, that will integrate the most up-to-date evidence and diverse perspectives. By involving expert development panel members and PFPs, the resulting CPG will be scientifically robust, patient-centered, and contextually relevant.

Voting on summary recommendations and question prioritization, along with a thorough literature review to develop EtD tables, ensures that the CPG addresses important clinical issues. This CPG is designed to promote optimal patient outcomes, making it valuable for clinicians, policymakers, patients and families.

## Data Availability

not applicable

## Acknowledgements

We would like to acknowledge the expert development panel members who reviewed the protocol: Gwenaelle De Clifford-Faugère, Pieter De Cock, Saskia deWildt, Dmytro Dmytriiev, Juliane Engel, Paola Claudia Fazio, Sylvia George, Isabelle Goyer, Anna Harðardóttir, Julia Harris, Klàra Horvàth, Erwin Ista, Santiago Mencía, Tuuli Metsvaht, Maria Cristina Mondardini, Mehdi Oualha, Marie-Hélène Perez, Krzysztof Pietrzkiewicz, Francesca Sperotto, Benjamin Wyness, and Nilufer Yalindag-Ozturk.

## Declarations

Data availability: Not applicable

Code availability: Not applicable

Authors’ contributions: IM: Conceptualization, Methodology, Writing – original draft. ACT: Conceptualization, Validation, Methodology, Writing – review & editing. CJ: Conceptualization, Validation, Methodology, Writing – review & editing. AA: Conceptualization, Validation, Methodology, Writing – review & editing. A-SR: Conceptualization, Methodology, Supervision, Validation, Writing – review & editing. ESPNIC PaDIS consortium: Conceptualization, Writing – review & editing. ESPNIC PaDIS consortium: Validation, Methodology, Writing – review & editing.

## Abbreviations

PICU: pediatric intensive care unit
IWS: iatrogenic withdrawal syndrome
ESPNIC: European Society of Pediatric and Neonatal Intensive Care
CPG: clinical practice guideline
GRADE: Grading of Recommendations Assessment, Development and Evaluation
EtD: evidence to decision
PFP: Patient and family partner
COI: conflict of interest
AGREE: Appraisal of Guidelines for Research and Evaluation
AGREE-REX: Appraisal of Guidelines for Research and Evaluation Recommendation Excellence
AMSTAR 2: A MeaSurement Tool to Assess systematic Reviews 2
SoF: Summary of findings

## Supporting information

**S1 File. Checklist for guideline protocol.**

**S2 File. Declaration of interest form and conflict of interest: Expert panel.**

**S3 File. <u>Information sheet: Patient and family partners.</u>**

**<u>S4 File.</u> Expression of interest form: Patient and family partners.**

**S5 File. Recommendation table.**

